# Unusual profile of germline genetic variants in unselected colorectal cancer patients from a high-prevalence region in Panama

**DOI:** 10.1101/2025.04.19.25326043

**Authors:** Iván Landires, José Pinto, Raúl Cumbrera, Alexandra Nieto, Gumercindo Pimentel-Peralta, Yennifer Alfaro, Virginia Núñez-Samudio

## Abstract

The profile of germline genetic variants among colorectal cancer patients in Panama has not yet been explored. We recruited 95 patients with colorectal cancer in an Oncology Reference Hospital Unit in the Azuero region of central Panama, which exhibits the highest prevalence of colorectal cancer in Panama. DNA analysis was performed with a panel of 113 genes with germline mutations for cancer (TruSight® Cancer Sequencing Panel from Illumina). Among the 95 cases, 10 pathogenic/likely pathogenic variants (P/LP) were identified in the MUTYH, TP53, CHEK2, PALB2, ATM and BARD1 genes, representing 10% of the total. The median age of colorectal cancer onset in variant-positive cases was 71 years. The variant 1103G>A (p.Gly368Asp) in MUTYH was the most prevalent. The variant at c.1675_1676delCAinsTG (p.Gln559Ter) in PALB2 is new and is reported for the first time in this study. The genes where variants were most frequently detected were MUTYH (5 patients) and CHEK2 (2 patients). It is striking that in Panama, in this first cohort of 95 patients studied for genetic variants of susceptibility to colorectal cancer, we did not find any patient with mutations in mismatch repair (MMR), which in other cohorts around the world are the most frequently mutated genes found. This atypical genetic profile of germline genetic variants could be related to the unique mestizo characteristics of the population living in Azuero in the central region of Panama and could explain, in part, the high prevalence of colorectal cancer in its inhabitants.

## Introduction

According to the World Health Organization (WHO), colorectal cancer is the third most common cancer and the second leading cause of cancer deaths worldwide (1). Incident cases of colorectal cancer more than doubled worldwide in the past three decades (2).

Panama is a country of 4.4 million inhabitants located in Latin America at the southern end of Central America. In Panama, cancer of all types is the second leading cause of death, and colorectal cancer ranks fourth in morbidity and mortality among all cancers at the national level. Interestingly, in Azuero, the central region of Panama, which includes the central provinces of Herrera, Los Santos, and the extreme south of the province of Veraguas, colorectal cancer ranks second in frequency and is also the leading cause of death from cancer-related causes (3). Due to this high frequency and mortality rate in Azuero, which is higher than the national rate and the highest in the country, it is necessary to carry out studies to identify why colorectal cancer is so highly prevalent in Azuero.

According to Panama’s 2023 census data, the country has 4,202,572 inhabitants, of which 31.7% is Afro-descendant, 17.2% is Amerindian and the remaining 51.2% is mestizo (4). The population of the Azuero region has ethnic and cultural characteristics that make it unique in the Republic of Panama. Currently, Azuero is considered one of Panama’s most Hispanicized regions due to the Spanish conquest and colonization. In addition, Azuero has been distinguished by its own culture that highlights Spanish and, to a lesser extent, Amerindian and African traditions, mixed in a singular way to give as a result the Azuero culture of today, which is unique in Panama (5). For this reason, we propose that certain genetic determinants of predisposition to colorectal cancer may be characteristic of the inhabitants of Azuero.

A 25 to 30% of cases of colorectal cancer are related to non-modifiable risk factors such as genetic factors, a personal history of polyps or adenoma, or a family history of colorectal cancer such as Lynch syndrome or familial adenomatous polyposis (2). Genetic studies of germline variants predisposing to colorectal cancer have identified that pathogenic and/or likely vpathogenic heterozygous variants (PVs/LPVs) in mismatch repair (MMR) genes are the most frequent genetic causes of colorectal cancer, which in turn are related to Lynch syndrome, also known as hereditary nonpolyposis colorectal cancer (HNPCC). MMR is essential for fixing DNA mismatches during DNA replication. When the MMR mechanism is defective, somatic mutational events accumulate in some genes containing tandemly repeated DNA motifs called microsatellites. Monoallelic APC PVs and biallelic inactivation of MUTYH are frequent causes of hereditary adenomatous polyposis and colorectal cancer (6, 7, 8).

In this study we present an unusual profile of germline genetic variants predisposing to colorectal cancer in which the genes MUTYH, CHEK2, TP53, PALB2, ATM and BARD1 predominate. This unique genetic profile could be related to the unique mestizo characteristics of the population living in Azuero in the central region of Panama and could explain, in part, the high prevalence of colorectal cancer in its inhabitants.

To the best of our knowledge, this is the first study to evaluate germline variants of genetic predisposition to colorectal cancer in patients from Panama.

## Materials and methods

We conducted a study of unselected colorectal cancer patients, diagnosed at any age, undergoing treatment and follow-up between March 2024 and March 2025 at the Unidad Oncológica de Azuero del Hospital Anita Moreno, the main referral hospital for cancer diagnosis and treatment in the central region of Panama.

To be enrolled in this study of germline genetic variants in patients diagnosed with colorectal cancer recruited without selection criteria, patients had to be at least 18 years of age, regardless of sex, mentally competent, and willing to sign informed consent documents. All patients in the study (n=95) were diagnosed with colorectal cancer at the Unidad Oncológica de Azuero del Hospital Anita Moreno. All patients included in this study voluntarily agreed to participate and gave written informed consent. Blood samples were collected with the approval of the Comité de Bioética de la Investigación del Hospital Chicho Fábrega, study number CBI-HRLCHF EC-CBIHRLCHF-2023-08102.

Each patient’s family history of cancer was collected using three-generation family trees. Ten millilitres (10mL) of venous blood was drawn from the forearm vein of consenting study participants (EDTA tetrasodium anticoagulant). Samples were preserved at 4^°^C briefly before genomic DNA extraction using the QIAamp® DNA Mini Kit (Qiagen, Hilden, Germany). DNA concentration was measured using Qubit 4TM Fluorometer (Thermo Fisher Scientific, USA). DNA samples with a concentration above 1.0 ug were used to prepare sequencing libraries using the TruSight Hereditary Cancer Panel to assess germline mutations across 113 genes for variant identification and were sequenced on a MiniSeq (Illumina, Inc, San Diego, CA) according to the manufacturer’s protocols.

All high-quality data were then mapped to the human genome assembly using the *bwa-mem* algorithm. The aligned files were processed with the Genome Analysis ToolKit (GATK) to recalibrate base quality, realign indels and remove duplicates. This was followed by SNP and INDEL discovery and genotyping according to GATK Best Practices recommendations. All variant calls underwent quality score recalibration and filtering to remove low-quality variants. Genetic data were analyzed using Golden Helix®’s SVS 8.8.3 (Bozeman MT, USA), and quality control was performed as recommended in several previous papers. Variants were filtered for missense or frameshift mutations, stop gains or losses, initiator codons, in-frame insertions or deletions and splice site alterations. Variants with a minor allele frequency < 0.25 were excluded from analysis. The predicted effect of variants was evaluated using ClinVar (9).

## Results

A total of 95 unselected colorectal cancer patients were tested for *cancer predisposition* gene variants using the TruSight Hereditary Cancer Panel. On average, 2.3 years had elapsed between the date of diagnosis and the date of interview and blood sample collection. The median age of the patients at diagnosis was 67 years and the median age at interview was 70 years. 7% of the patients were diagnosed before the age of 50 and 28.4% were diagnosed before the age of 60.

A P/LP variant was found in 10 of 95 (10.5%) patients; 4 had a P/LP variant in *MUTYH* (4.2%), 2 had a P/LP variant in *CHEK2* (2%), 1 had two P/LP variants in *TP53* (1%), 1 had a P/LP variant in *ATM* (1%), 1 had a P/LP variant in *BARD1* (1%) and 1 had a P/LP variant in *PALB2* (1%) (See Table 1). 1 patient had 2 different P/LP variants in *MUTYH* (p.Gly368Asp and p.Tyr151Cys). The p.Arg81Trp variant in MUTYH was identified in another patient. The p.Gly368Asp variant in MUTYH was the most prevalent with 3 of 4 patients with variants in MUTYH. The second gene with the highest frequency of P/LP variants was CHEK2, with two patients with variants p.Glu273Lys and p.Arg117Gly with ages ranges at onset of 41-45 and 61-65 years, both with no history of first-degree relatives but with second-degree relatives affected. A male patient with 2 variants in TP53 was also identified: c.783-1G>A affecting splicing and a missense variant p.His179Gln. A patient with a p.Ile1332fs frameshift variant in ATM and another patient with the p.Glu59fs variant in BARD1 was identified. The p.Gln559Ter variant in PALB2 is new and is reported for the first time in this study in a young patient with age range at onset between 31-35 years and with a history of first- and second-degree relatives affected by cancer.

**Table 1.** MC : Molecular Consequence, M : Missense, N : Nosense, F : Frameshift, ACMG : ACMG Clasification, LP : Likely Pathogenic, P :Pathogenic

| ID | Gene | Variant | M<br>C | rs<br>identifie<br>r | HGVS nomenclature<br>(GRCh38.p14) | AC<br>M<br>G | Gender | Age range<br>at diagnosis<br>(years) | First- and second-degree family<br>history of cancer |
| --- | --- | --- | --- | --- | --- | --- | --- | --- | --- |
| CC_14 | MUTYH | c.1103G>A<br>(p.Gly368Asp) | M | rs360539<br>93 | NC_000001.11:g.45331556<br>C>T | LP | Male | 71-75 | One parent with colon cancer and the<br>another parent with pancreas cancer, 2<br>siblings with colon cancer |
| CC_14 | MUTYH | c.452A>G<br>(p.Tyr151Cys) | M | rs346123<br>42 | NC_000001.11:g.45332803<br>T>C | LP | *Patient<br>CC_14 as<br>above | *Patient<br>CC_14 as<br>above | *Same patient ID: CC_14 as above |
| CC_47 | MUTYH | c.1103G>A<br>(p.Gly368Asp) | M | rs360539<br>93 | NC_000001.11:g.45331556<br>C>T | LP | Male | 86-90 | Parent with colon cancer, sibling with<br>prostate cancer |
| CC_69 | MUTYH | c.1103G>A<br>(p.Gly368Asp) | M | rs360539<br>93 | NC_000001.11:g.45331556<br>C>T | LP | Female | 71-75 | None |
| CC_03 | MUTYH | c.241C>T<br>(p.Arg81Trp) | M | rs765123<br>255 | NC_000001.11:g.45333436<br>G>A | LP | Female | 61-65 | Parent with colon cancer, sibling with<br>colon cancer |
| CC_50 | TP53 | c.783-1G>A | A<br>S | rs155552<br>5367 | NC_000017.11:g.7673838C<br>>T | LP | Male | 81-85 | None |
| CC_50 | TP53 | c.537T>A<br>(p.His179Gln) | M | rs876660<br>821 | NC_000017.11:g.7675075A<br>>T | P | *Patient<br>CC_50 as<br>above | *Patient<br>CC_50 as<br>above | *Same patient ID: CC_50 as above |
| CC_28 | CHEK2 | c.817G>A<br>(p.Glu273Lys) | M | rs587782<br>152 | NC_000022.11:g.28710035<br>C>T | P | Male | 41-45 | None |
| CC_54 | CHEK2 | c.349A>G<br>(p.Arg117Gly) | M | rs289099<br>82 | NC_000022.11:g.28725338<br>T>C | LP | Female | 61-65 | None |
| CC_48 | PALB2 | c.1675_1676del<br>CAinsTG<br>(p.Gln559Ter) | N | rs587780<br>206 | NC_000016.10:g.23634870_<br>23634871inv | P | Female | 31-35 | Parent with colon cancer, 2 siblings with<br>colon cancer |
| CC_58 | ATM | c.3994del<br>(p.Ile1332fs) | F | N/A | NC_000011.10:g.108287600<br>del | P | Male | 76-80 | None |
| CC_60 | BARD1 | c.176_177del<br>(p.Glu59fs) | F | rs105751<br>7589 | NC_000002.12:g.214797104<br>_214797105del | LP | Female | 71-75 | Parent with prostate cancer |

2 of all 10 P/LP variant positive patients were diagnosed under age 50 years (20%) whereas among those patients negative for P/LP variants only 3 of 85 patients were diagnose under 50 years old (3.5%). The prevalence of P/LP variants was 33% among those diagnosed under age 50 years, and 8% for participants over the age of 50 years.

5 of the 10 patients with a P/LP variant had a first-degree relative affected with cancer (50%) (See Table 1), whereas only 33 of 85 patients without a P/LP variant identified had a first-degree relative affected with cancer (38%). Other cancers in the ten families with a P/LP variant included pancreas cancer (3 cases), breast cancer (1 case), prostate cancer (1 case) and liver cancer (1 case).

## Discussion

Germline genetic variants have been studied in 95 patients with colorectal cancer from a highly prevalent region in Panama. In 10.5% of the total patients, P/LP with an atypical profile were identified, since 4% and 2% corresponded to patients with variants in the MUTYH and CHEK2 genes, respectively. No patients were found to have germline variants in the MMH genes, which have been reported in other studies as the most frequent in colorectal cancer.

To our knowledge, this study presents the first evaluation of germline genetic variants associated with colorectal cancer in the Panamanian population. It has been carried out in Azuero, a central region of Panama, where the prevalence of the disease is the highest in the country. In Azuero, the Spanish contribution to mestizaje is high in the population, in contrast to the rest of the country, where the contribution of Amerindian and African descendants is higher (5) which could explain, in part, the existence of genetic determinants specific to this population that explain the high rates of colorectal cancer in the region. The results show that approximately 10.5% of patients diagnosed with colorectal cancer in Panama exhibit pathogenic/ likely pathogenic (P/LP) variants in cancer predisposition genes, which highlights the importance of investigating the genetic basis of this disease in our population.

The most frequent finding was that of P/LP variants in the MUTYH gene identified in 4 patients, the most prevalent being the p.Gly368Asp variant, which was found in 2 patients in monoallelic presentation, while in a third patient this p.Gly368Asp variant and the p.Tyr151Cys variant were found in probable biallelic presentation. The very rare p.Arg81Trp variant in MUTYH was found in monoallelic presentation in a fourth patient. The finding of the p.Gly368Asp variant in MUTYH as the most frequent variant is important because it is consistent with previous studies that have found this variant to be important in the Eurasian population. The p.Tyr151Cys variant has also been reported to be significant in frequency among patients with MUTYH mutations and colorectal cancer (10). Of these 4 patients with Ps/LPs variants in MUTYH, in 3 of them, a history of first-degree relatives affected by cancer was identified. Up to 2% of people of Northern European descent have been found to carry a pathogenic variant in MUTYH (10). In our series of unselected patients up to 4% were carriers of Ps/LPs variants in MUTYH, which consolidates the importance of this marker in the germline genetic evaluation of patients with colorectal cancer in Panama. The fact that one of the patients presented 2 variants in MUTYH in probable biallelic presentation, while the other 3 patients presented 1 single variant in MUTYH (monoallelic presentation), could suggest that these monoallelic variants are of high penetrance in our population. Previous studies have shown that MUTYH-associated polyposis (MAP) and colorectal cancer are strongly associated with bi-allelic germline pathogenic variants, whereas a two-fold increased risk has been observed in monoallelic carriers with a family history of cancer. The results of the present study are compatible with a recent study of germline variants in the MUTYH gene in Latin American patients with colorectal cancer, where it was found that the most frequent variant was p.Gly368Asp gene (10). It is important to emphasize that in this study, 2 of 3 patients with monoallelic presentation had a history of cancer in first-degree relatives, which points to the importance of studying the family for genetic counseling and cancer prevention interventions in relatives who test positive for these variants, even in monoallelic presentation.

The second most frequent gene with P/LP variants was CHEK2 with 2 patients (2%). We identified one patient with the p.Glu273Lys variant whose age of disease onset was early in a range age at onset of 41-45 years and another patient with the p.Arg117Gly variant with age range of onset of 61-65 years, both with no history of first-degree relatives affected by cancer but with second-degree relatives affected by cancer. In our sample, the age of onset of colorectal cancer was relatively young in patients with Ps/LPs variants in CHEK2, which suggests that this gene could be involved in cases of early onset of colorectal cancer in our population. It should be added that, recently, the role of the CHEK2 gene in predisposition to colorectal cancer has been the subject of controversy. Although some studies suggest a slight association between certain CHEK2 mutations and an increased risk of colorectal cancer, it should be added that the role of the CHEK2 gene in predisposition to colorectal cancer has recently been controversial (11), as others indicate that this relationship may not be significant. For example, the 2024 National Comprehensive Cancer Network (NCCN) guidelines indicate that there is no increased risk of colorectal cancer associated with variants in CHEK2, recommending consideration of other factors to assess individual risk, but, the fact that in our cohort both patients with P/LP variants in CHEK2 are younger than the mean age at diagnosis in this cohort (67 years) and that both have a family history suggests that in our population CHEK2 may play a role in the predisposition to colorectal cancer in the Panamanian population (12).

An old male patient with an age range at onset between 81-85 years with no family history of cancer had two variants in TP53: c.783-1G>A (likely pathogenic) and p.His179Gln (pathogenic), which were predicted to affect splicing and missense, respectively. Mutations in TP53 cause Li-Faumeni syndrome, a disease that predisposes to multiple familial cancers (13). In several cohorts, TP53 has been found to be a predisposition gene for colorectal cancer (14). The absence of a family history of cancer and the advanced age of the patient suggest incomplete and reduced penetrance of these variants or the influence of modifying factors that have delayed the age of colorectal cancer onset in this patient. This points to the difficulty of genotype-phenotype correlation in TP53 mutations and the need to take into account other additional factors in the assessment of oncologic risk.

A male patient with an age range at onset between 76-80 years with no family history presented the pathogenic variant p.Ile1332fs in ATM, predicted to introduce a frameshift, positioning himself also as a case of interest in the evaluation of genetic predisposition to colorectal cancer. The ataxia telangiectasia (ATM) mutation is widely documented for its involvement in the repair of DNA double-strand breaks within the DNA damage response (DDR) pathway, as pathogenic mutations in this gene can compromise genomic stability and increase susceptibility to various malignancies (15). The p.Ile1332fs mutation introduces a change in the reading frame that leads to nonsense-mediated mRNA decay (NMD), which interferes with its normal function in DNA repair. It can be hypothesized that frameshift variants such as the one carried by this patient may be more prevalent than expected than other types of variants in ATM. The latter could explain the development of the colorectal cancer phenotype in this patient with no significant family history and with a probable de novo variant in ATM.

An LP p.Glu59fs variant in the BARD1 gene was reported in a female patient with an age range of onset of colorectal cancer between 71-75 years and a first- and second-degree family history of neoplasms other than colorectal cancer. BARD1 AND BRCA1 form a heterodimer with multiple tumor suppressor functions related to DNA repair and apoptosis. Pathogenic variants in BARD1 have been shown to have moderate penetrance in triple-negative breast cancer (16). Recent studies have shown that pathogenic variants in BARD1 may also play a role in colorectal cancer in families with a positive family history of the disease (17). The patient reported in this study exhibits a family history of a first-degree relative with prostate cancer and with renal cancer, and third-degree with colorectal cancer, which underlines that the variant reported in this study would be in the causality of colorectal cancer in the patient and neoplasms in the family.

An unexpected and novel heterozygous variant in PALB2, p.Gln559Ter, has been found (rs587780206). To the best of our knowledge, this nonsense variant has never been reported in disease-related databases, including the Human Gene Mutation Database (HGMD), ClinVar or population databases, including gnomAD SVs v2.1. The finding of the variant chr16-23634870-TG-CA (PALB2:p.Gln559Ter) in a patient with colorectal cancer, who also presents a family history of the same disease in first and second degree, suggests as a hypothesis a possible implication of this genetic alteration in the predisposition to colorectal cancer. This variant is characterized by a nonsense-type mutation (null variant) that introduces a premature stop codon in exon 4 of 13 of the PALB2 gene, leading to NMD. The variant in question is a 2 base pair substitution leading to the change of TG for CA at chromosomal positions 1675 and 1676 (see Table 1) which shares position with a variant already known to be pathogenic with a single change of G for A at position 1675 leading to the same effect in translation: p.Gln559Ter (rs1555461154). The American College of Medical Genetics (ACMG) criteria for the novel variant reported in this study are PVS1, PM2, which classify it as pathogenic. Although mutations in PALB2 have primarily been associated with an increased risk of other cancers, such as breast cancer, recent studies have begun to explore its role in hereditary colorectal cancer (18). The lack of a previous record of the chr16-23634870-TG-CA variant in genetic databases, as well as its non-appearance in the existing scientific literature, indicates that this is a novel variant. This novelty, together with the clinical presentation of the patient, highlights the need to include this variant in future scientific studies to evaluate its relevance in predisposition to colorectal cancer.

Notably, the data from this study showed a higher frequency of P/LP variants in patients diagnosed before the age of 50 years (33% vs. 8% in those older than 50 years). In addition, 20% of patients with P/LP variants were diagnosed before the age of 50, whereas only 3.5% of patients without P/LP variants were diagnosed before the age of 50. This is consistent with the literature showing that patients diagnosed at a younger age are more likely to carry predisposing genetic variants, strongly suggesting the need for earlier genetic surveillance in younger individuals (18).

In this study, the presence of a family history of cancer in the families of patients with P/LP variants also indicates a possible hereditary predisposition to colorectal cancer. Fifty percent of patients with P/LP variants reported having a first-degree relative with cancer, compared to 38% in the group without genetic variants. This is consistent with the idea that a family history of cancer should be an important factor in assessing colorectal cancer risk (19).

The results of this study are consistent with those reported in diverse populations around the world, where a frequency of 5 to 15% of germline genetic variants have been found in patients with colorectal cancer. However, the specific variants may vary depending on the ethnic characteristics of the population studied. This highlights the need for further studies in specific Latin American populations, where specific genetic characteristics may influence the results (10).

One of the most remarkable and atypical findings of this study is the absence of pathogenic variants in key mismatch repair (MMR) genes, such as MLH1, MSH2, MSH6 and PMS2, which are the most common in other international cohorts of colorectal cancer patients, representing an important difference from what is observed in other populations. Colorectal cancer associated with predisposing pathogenic variants in the MMR genes is one of the most common forms of hereditary colorectal cancer and is responsible for Lynch syndrome, which affects approximately 2-3% of all colorectal cancer cases in various cohorts worldwide, including Latin American cohorts (20). This lack of variants in the MMR genes in the Panamanian population would have implications. First, it suggests that defects in DNA repair may not be a predisposing factor for colorectal cancer in Panama as they are in other populations. This discrepancy raises an important question about the specific genetic mechanisms that might be involved in the development of colorectal cancer in Panama and indicates that other genetic factors, such as the variants found in this study, are the most important in our population. The absence of variants in the MMR genes could indicate that diagnostic protocols and prevention strategies based on genetic testing of these genes are not effective in this specific population. Therefore, it would be beneficial to include in the germline variant evaluation other genes such as MUTYH, CHEK2, TP53, ATM, BARD1, and PALB2, which are relevant in our cohort and which are atypical in other populations worldwide. This atypical genetic profile of germline genetic variants may be related to the unique mestizo characteristics of the population of Azuero, central Panama, and may partially explain the high prevalence of colorectal cancer among its inhabitants.

Given the high frequency of colorectal cancer in Azuero, more extensive research with larger cohorts is needed to better assess the role of other genetic variants and environmental factors that may contribute to this high prevalence. The creation of specific genetic databases for the Panamanian population as a whole, but especially for populations with a high burden of colorectal cancer, will be critical for the development of personalized prevention and treatment strategies.

In addition, this study highlights the need for further research in Latin American populations where the particular genetic background may influence the prevalence of colorectal cancer predisposing variants. The identification of new or rare genetic variants in genes such as PALB2 raises the need to further investigate the role of new variants and predisposition genes for colorectal cancer.

It is important to recognize that this study has limitations due to the relatively small sample size and the lack of a well-defined control group. Also, this study was localized to the Azuero region, where colorectal cancer rates are the highest in Panama, but it should be considered that the genetic predispositions to colorectal cancer in Azuero are not necessarily the same as for the rest of the Panamanian population or for other Latin American populations. In addition, although the panel of genes used is broad, there may be other relevant genetic variants that were not evaluated in this study. Therefore, future research including a wider range of genes and a larger number of patients from different regions should be conducted to deepen the relationship between genetic variants and colorectal cancer in the Panamanian and Latin American populations.

Finally, future research in understudied populations must continue to evaluate the relationship between genetic variants and colorectal cancer predisposition, as well as their potential impact on treatment response.

## Data Availability

All data generated or analysed during this study are
included in this published article

## Acknowledgments

Iván Landires and Virginia Núñez-Samudio are members of the Sistema Nacional de Investigación (SNI), which is supported by Panama’s Secretaría Nacional de Ciencia, Tecnología e Innovación (SENACYT).

## Authors’ contributions

Conceptualization, I.L., J.P., and V. N-S.; methodology, I.L., J.P., R.C., A.N., G.P.P., Y.A., and V. N-S.; software, I.L., J.P., R.C., and V. N-S.; validation, I.L., J.P., R.C., and V. N-S.; formal analysis, I.L., J.P., R.C., A.N., G.P.P., Y.A., and V. N-S.; investigation, I.L., J.P., R.C., A.N., G.P.P., Y.A., and V. N-S.; resources, I.L., J.P., and V. N-S.; data curation, I.L., J.P., R.C., A.N., and V. N-S.; writing—original draft preparation, I.L., and V. N-S.; writing—review and editing, I.L., J.P., R.C., A.N., G.P.P., Y.A., and V. N-S.; visualization, I.L., J.P., R.C., A.N., G.P.P., Y.A., and V. N-S.; supervision, I.L., J.P., R.C., and N-S.; project administration, I.L., and V.N-S.; funding acquisition, I.L., J.P., Y.A., and V. N-S.; All authors have read and agreed to the published version of the manuscript.

## Availability of data and materials

All data generated or analysed during this study are included in this published article.

## Ethics approval and consent to participate

The study was conducted according to the guidelines of the Declaration of Helsinki, and approved by the Comité de Bioética de la Investigación del Hospital Chicho Fábrega, study number CBI-HRLCHF EC-CBIHRLCHF-2023-08102. No further administrative permissions were needed to access the raw data used in this study. The data used in this study were anonymized before use.

